# Modeling the COVID-19 outbreak in the United States

**DOI:** 10.1101/2020.04.30.20086884

**Authors:** Charit Samyak Narayanan

**Author notes:** These authors contributed equally to this work. Mission San Jose High School, 41717 Palm Ave, Fremont, California, United States of America.

## Abstract

The COVID-19 contagion has developed at an alarming rate in the US and as of April 24, 2020, tens of thousands of people have already died from the disease. In the event of an outbreak like such, forecasting the extent of the mortality that will occur is crucial to aid the implementation of effective interventions. Mortality depends on two factors: the case fatality rate and the case incidence. We combine a cohort-based model that determines case fatality rates along with a modified logistic model that evaluates the case incidence to determine the number of deaths in all the US states over time; the model is also able to include the impact of interventions. Both models yield exceptional goodness-of-fit. The model predicted a range of death outcomes (79k to 246k) all of which are considerably greater than the figures presented in mainstream media. This model can be used more effectively than current models to estimate the number of deaths during an outbreak, allowing for better planning.

## Introduction

The first case of coronavirus disease 2019, or COVID-19, a respiratory infection caused by severe acute respiratory syndrome coronavirus 2 (SARS-CoV-2), was first identified in Wuhan, China in late 2019 [1]. Subsequently, the outbreak has spread to 212 [2] countries, including the United States, where the first case of COVID-19 was detected in Washington state on January 20, 2020 [3]. As of April 24, 2020, the U.S has reported 830053 cases and 42311 deaths [4].

As the pandemic progresses, determining its prognosis is essential to inform the adoption of adequate mitigation efforts. Many have attempted to forecast the trajectory of the epidemic in the United States, and at the forefront is the White House Coronavirus (COVID-19) Task Force [5], led by Dr. Anthony Fauci [6]. On March 29, Dr. Fauci suggested that the US would likely face 100,000 to 200,000 deaths, with millions of cases [7]. However, on April 9 he said the estimate had been revised down to 60,000 [8]. Moreover, a model by the University of Washington, closely followed by the White House, projects 67,000 deaths as of April 24, 2020 [9], in line with Dr. Fauci’s statement [8].

It is well-known that there is a wide variance in the sizes of outbreaks between states as well as the resulting incidence of deaths. The primary reason for the variance is the uncertainty in the prediction of infections and the fatality rate of infected individuals. In this paper, we forecast the final number of cumulative deaths in each state using improved models to determine both the cumulative case incidence and the case fatality rate. We calculated the case fatality rate (CFR) for each state using a cohort-based approach, which has demonstrated greater accuracy than traditional methods [10]. Additionally, the number of cumulative cases in each state is predicted using a modified logistic model. Combining the two, we are able to forecast the number of cumulative deaths by state. Additionally, we analyze the drivers of deaths and discuss implications on policy formation.

## Methods

### Data sources

The primary data for this study is publicly available and was obtained from the Center for Systems Science and Engineering (CSSE) at Johns Hopkins University [11]. We obtained the data pertaining to daily new cases/deaths and cumulative cases/deaths for the period of January 22, 2020, to April 24, 2020. In addition, we obtained US state population data from the 2010 United States Census Bureau [12]. The number of deaths is a product of the case fatality rate (CFR) and the population confirmed to have been infected [13]. Therefore, in order to determine cumulative fatalities due to COVID-19, it is necessary to first predict the CFR and the number of cases. CFR, case incidence, and deaths were evaluated for all fifty states as well as the District of Columbia.

### Calculating CFR

There are three principal measures of disease lethality: the case fatality rate (CFR), infection fatality rate (IFR) and mortality rate (MR). The mortality rate is represented by the proportion of cumulative deaths to the total at-risk population. This is ultimately indicative of the probability of any individual’s mortality among the total population. The CFR uses the same numerator (cumulative deaths) but instead divides it by the number of cumulative confirmed cases. The case fatality [14] rate is the proportion of individuals who die from a disease among all individuals diagnosed with the disease within a specified timeframe [15]. That is, it reveals the percentage of individuals that die among all individuals who test positive for the disease. The IFR is similar to the CFR, except it represents the ratio of deaths to the total number of people who are infected; it accounts for all infected individuals regardless of whether their disease is reported or not. In an ideal scenario, where zero individuals with COVID-19 went unnoticed, and surveillance was faultless, the IFR and CFR would be equivalent. However, this is not truly plausible; testing is limited, asymptomatic infections commonly are not surveilled, and not all instances of the disease are accounted for in reality. As a result, the CFR that we calculate will be much higher than the IFR and the mortality rate.

In this paper, we use a logistical function [10] to describe the exponential growth and subsequent flattening of COVID-19 CFR. The CFR depends on three parameters: the final CFR (L), the CFR growth rate (k), and the onset-to-death interval (*t*_0_) and is expressed as:

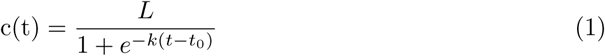

Using this model, we calculate the number of deaths each day for each cohort or group of individuals infected on the same day. Next, we build an objective function that minimizes the root mean square error between the actual and predicted values of cumulative deaths. We ran 125,000 simulations, using numerous values of the onset-to-death interval, the CFR, and the CFR growth rate. The CFR was kept in the range of 0.5% to 20%, the slope was kept in the range of 0.005 and 0.7, and the onset-to-death interval bounded between 0 and 60 days. We assigned these bounds because after in-depth explorations of the model, we were convinced the solutions would be within these parameters. We then identify the model parameters that best fit the data (top 1% of the best-fit RMSE). With a kernel density distribution of case fatality rates, we determined the low CFR (the lowest value regardless of its frequency); the mode CFR (the most probable CFR); and the high CFR (the highest value regardless of its frequency).

### Calculating Cumulative Cases

Most methods for calculating the spread of infections use a form of logistic or Susceptible-Infectious-Recovered (SIR) model [16] [17] [18]. While these models perform effectively when predicting the spread of infections in the absence of interventions, they are unable to model the outbreak (without additional modifications) under conditions of mitigation efforts such as shelter-in-place orders. Common to all methods is the incorporation of the growth rate. In the SIR model, this is *R*_0_, the transmission rate– given that the population lacks immunity and there are no deliberate interventions to impede disease transmission. The number of infections will continuously rise in a population if *R*_0_ > 1, will remain steady if *R*_0_ = 1, and will decrease if *R*_0_ < 1. To explicitly account for the impact of mitigation efforts, models must support gradual changes in the shape of the case growth rate.

The logistic model forecasts the slow initial rise, exponential growth, and eventual decay of cumulative cases, but cannot account for the changes that result from interventions. Interventions may drastically impact the rate of transmission, thus it is necessary to adapt the model to suit these conditions.

Therefore, we have adapted the logistic model to include the change in the rate of infection to include the effects of interventions such as social distancing. The original logistic model total that determines the total number of cases depends on three parameters: the terminal number of cumulative cases (C), the CFR growth rate (r), and the days to the inflection point (*t_i_*). The inflection point indicates the day at which the number of daily cases reaches its maximum. This function that describes the change in case incidence*i*(*t*) over time can be expressed as:

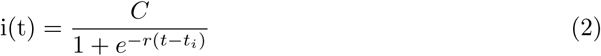

The modified logistic model has five parameters; however, the terminal number of cumulative cases (C) and inflection point (t0) remain unchanged. The set of equations that describe the incidence using the modified logistic model are:

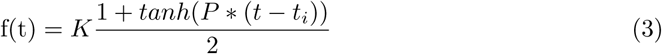

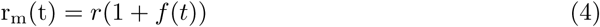

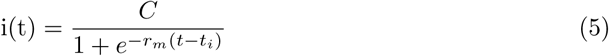

The parameter *r_m_* is the modified growth function that changes over time, and f is a smoothing function that determines how quickly the rate diverges around the point of inflection, as well as the magnitude of the transformation. Using this model, we calculated the number of cases for each state.

Next, we built an objective function that minimizes the root mean square error between the actual and predicted values of cumulative cases, and ran numerous simulations by varying the four parameters. We held P constant at 0.1 in all our simulations. We ran 375,000 simulations, using numerous values of the days to inflection: t0, the terminal number of cases, C, the growth rate, r, and the growth rate multiplier, K. The terminal number of cases was kept in the range of 0.01% to 3% of the each state’s penetration, the growth rate was kept in the range of 0.01 and 0.3, and the number of days to inflection was bounded between 10 and 50 days. The growth rate multiplier was kept in the range between 0 and 40%.

From these simulations, we identify the set of parameters that returns the lowest error. Finally, we forecasted the number of cases up to September 10, 2020. With a kernel density distribution of case incidence possibilities, we determined the mode number of cases (the most probable value of cumulative cases) and the 95% percent confidence interval range of case incidence for each state.

### Calculating Cumulative Cases

The cumulative mortality is the product of the case fatality rate and the cumulative case incidence. With the low, mode, and high values of both CFR and cumulative case incidence, we evaluate the nine possible death tolls for each state by finding the product of each CFR value and each value of the cumulative case incidence. To determine cumulative mortality on the national scale, we add up the respective cells for all states.

## Results

### Predicting case incidence

We calculated the case incidence for each jurisdiction. Fig 1 shows the goodness-of-fit between the forecasted cases and the true number of cases for New York. The two sets of figures show the cumulative case incidence and new case incidence. It demonstrates that there is an excellent fit for both new and cumulative cases. We calculated the *R*^2^ for all the states and the fit was excellent (greater than 98% for all the states) for all states indicating that the modified logistical function does a great job of modeling the transition after the intervention. More information regarding the model’s goodness-of-fit for the states is provided in the supplemental information (see S1 Table).

**Fig 1.**
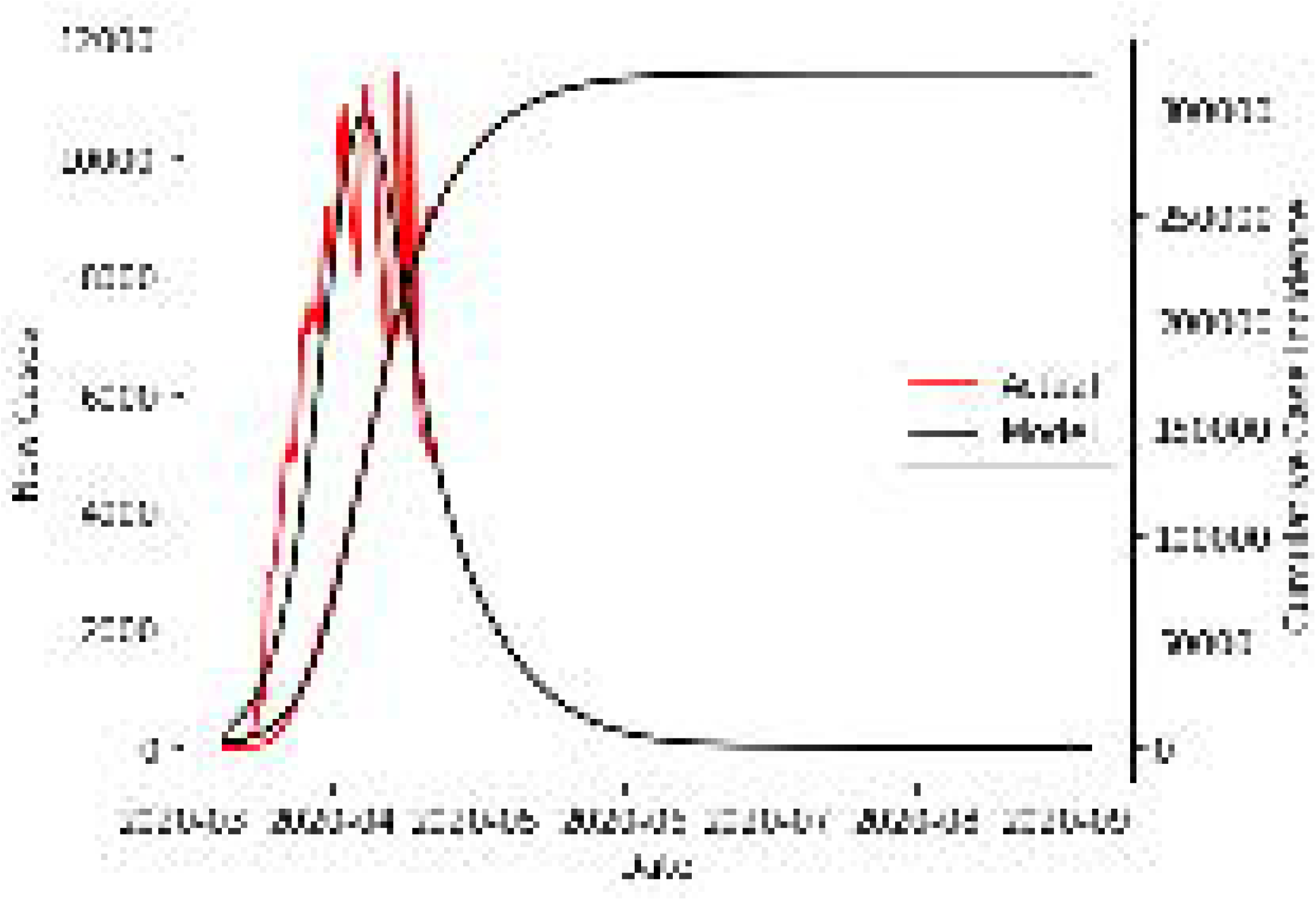
Comparison of modeled and actual number of cases for New York. The goodness-of-fit between the forecasted cases and the true number of cases for New York. Used data from March 10 through April 24.

Fig 2 displays the range of the forecasted total case incidence for the top states that contribute 90% of all the deaths. The vast majority of the states will experience less than 50000 cases (see S2 Table). However, New York is a substantial outlier: the model predicts 500,000 cases for the mode case. All the numbers we will quote in the rest of the document will be the mode case (unless otherwise specified), as it has the highest likelihood. Even if the best-case scenario transpires, its case incidence will probably exceed the incidence of any other state by over 200000.

**Fig 2.**
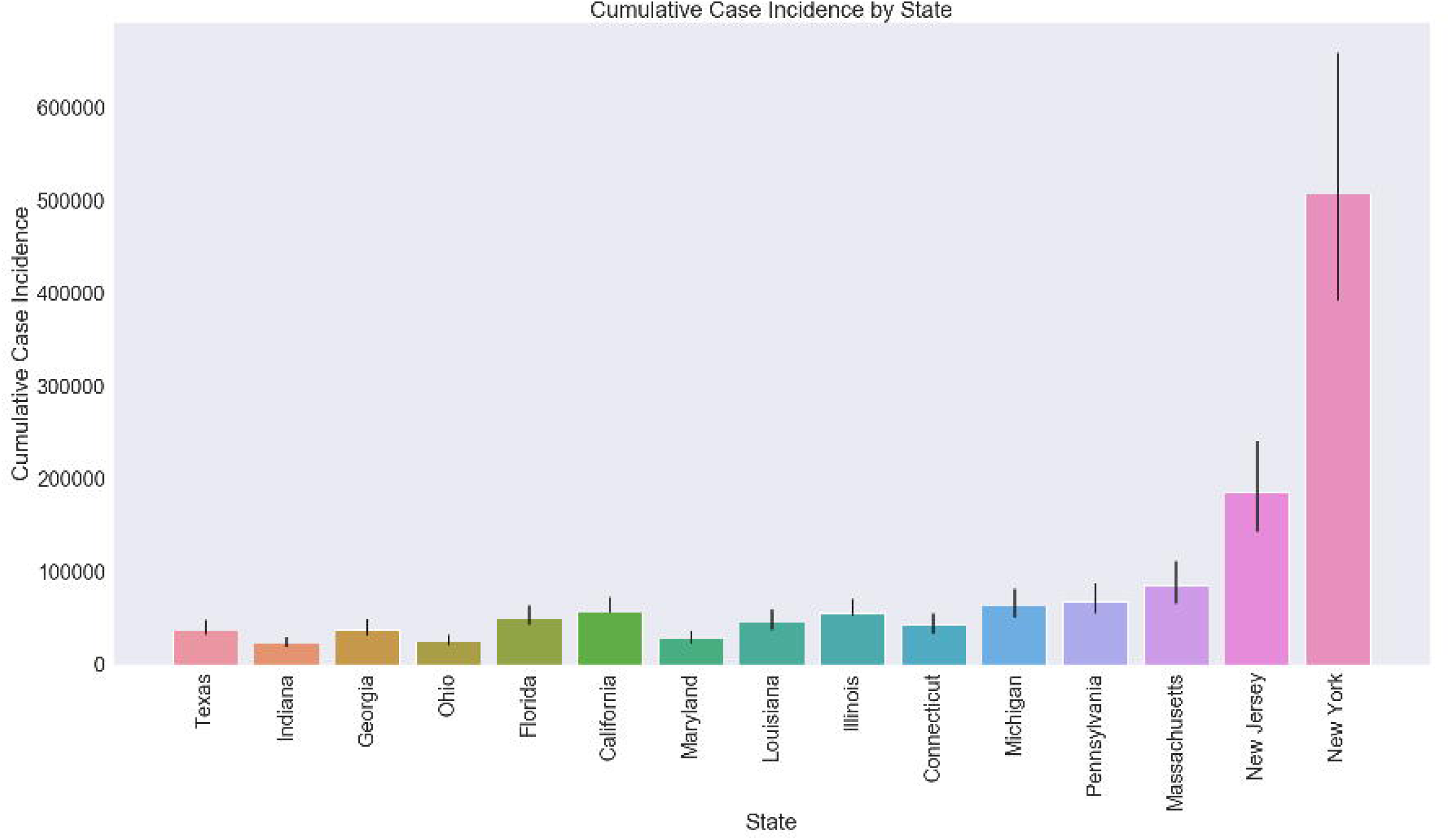
Forecasted case incidence for the top US states. The range of forecasted total case incidence for the top states that contribute 90% of all the deaths. The low, mode, and high cases are displayed. The low and high cases are determined as the low and high end of the 95% confidence interval.

Next, we evaluated the forecasted case incidence for the entire United States (Fig 3). The total number of cases predicted is above 1.2M, and the number of new daily cases peaks at more than 35000. As New York and New Jersey contribute significantly toward the overall case incidence, the United States peak daily cases is strongly dependent on the peak of these two states.

**Fig 3.**
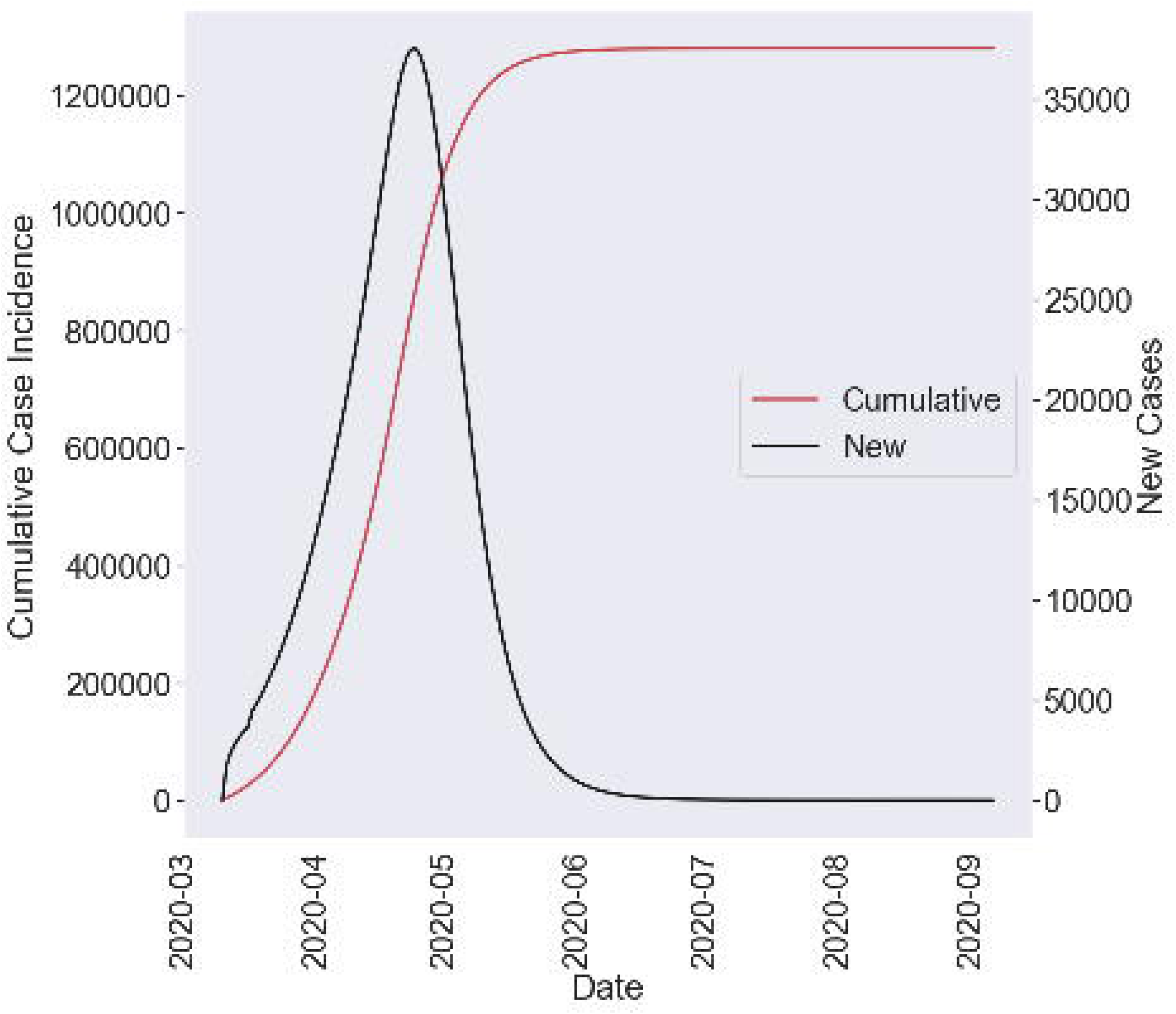
Forecasted Case incidence for US over time. The forecasted case incidence of the United States over time. This shows both the new and cumulative number of cases.

In order to understand whether the different states have a case incidence proportionate to their populations, we calculated the discrepancy between the projected cases per capita for individual states and U.S average projected cumulative cases per capita (Fig 4). New York, New Jersey, Massachusetts, Connecticut, and Louisiana have much higher case incidence per capita. This means that the disease affected these areas disproportionately. In contrast, the states of California, Texas, Florida, and Ohio did much better in controlling the spread of the infection.

**Fig 4.**
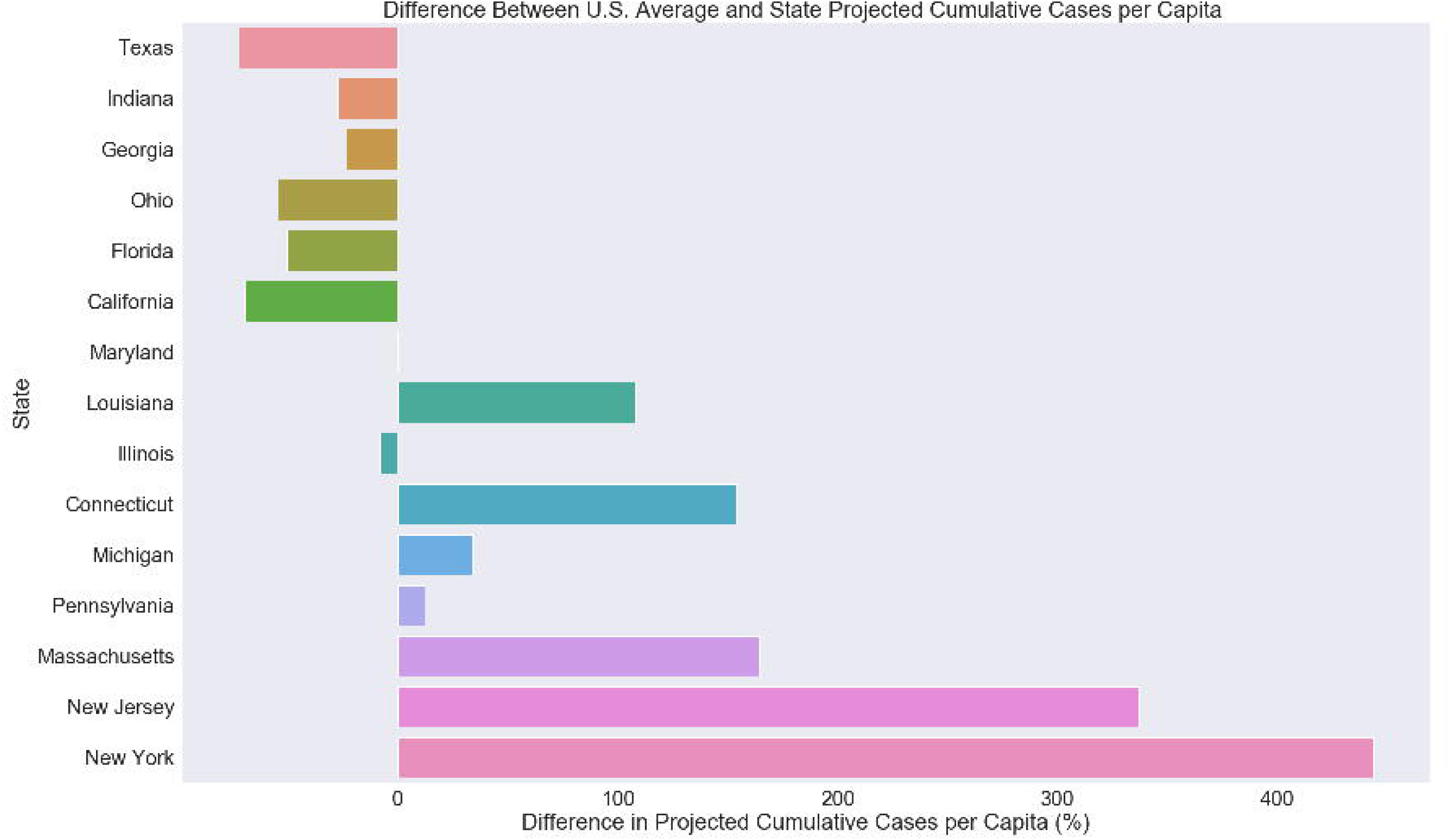
Difference in forecasted case incidence between states. Difference in forecasted case incidence between the top states that contributed 90% of all the deaths. This was calculated by subtracting the difference between projected cases per capita for individual states and the U.S average projected cumulative cases per capita.

### Predicting Case Fatality Rates

We previously calculated the case fatality rates for Hubei province and showed that the goodness-of-fit was excellent [10]. We used the same methodology and calculated the CFR for all the states. The model is able to fit the data extremely well showing that both the model and the methodology are sound. We provide the *R*^2^ value for all the states in the supplemental information (see S1 Table).

We calculated the range of final case fatality rates for each state (Fig 5). The CFR for most states is between 5% and 10%. Compared to the case incidence, case fatality rates have far less variability. Massachusetts, Connecticut, New York, and Maryland have relatively higher CFR’s. In contrast, Texas, California and Georgia have much lower CFR’s. We also provide supplemental information for all state (see S3 Table).

**Fig 5.**
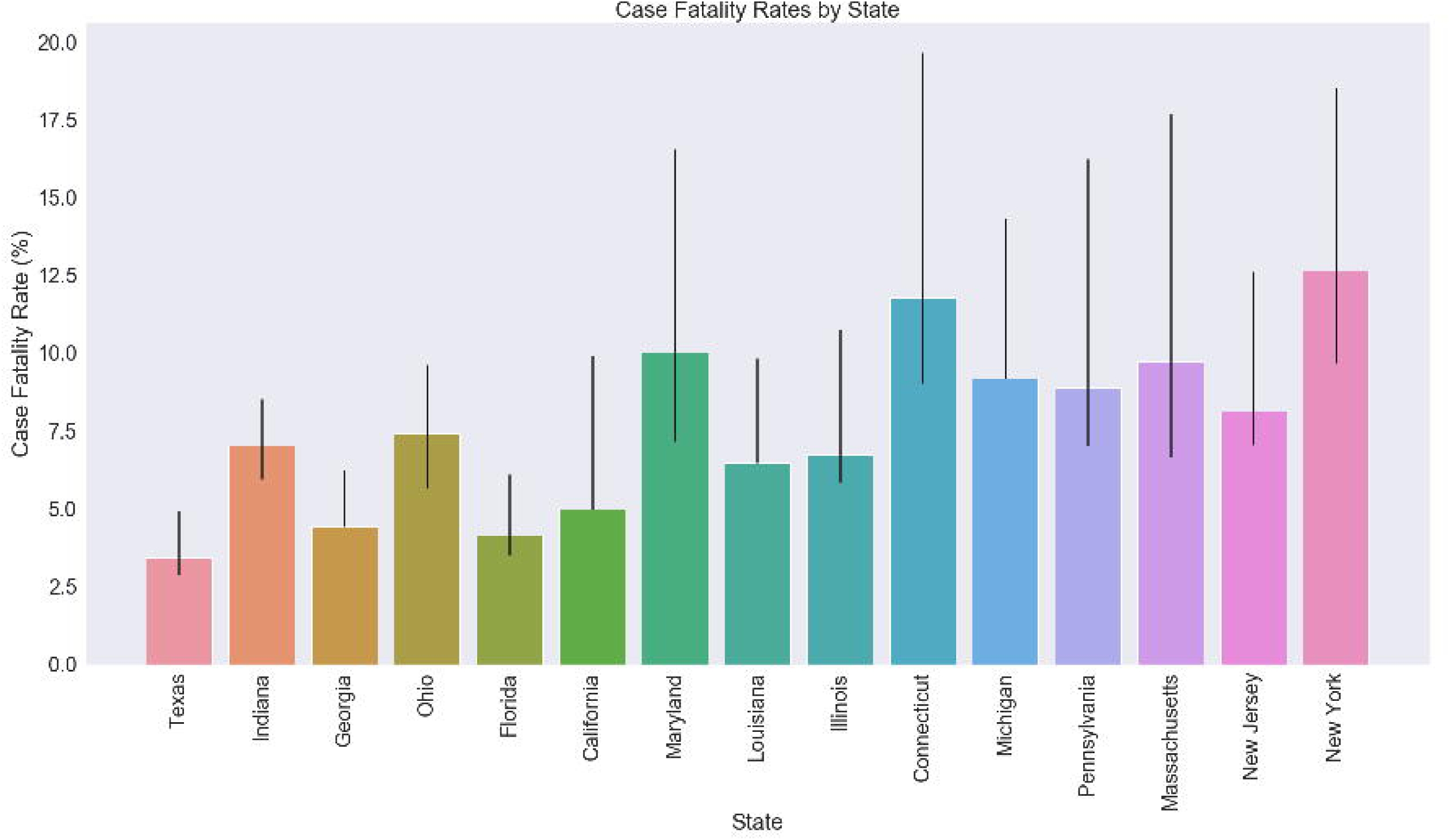
Forecasted case fatality rates for the top US states. The range of forecasted total fatality rates for the top states that contribute 90% of all the deaths. The low, mode, and high cases are displayed. The low and high cases are determined as the low and high end of the 95% confidence interval.

In order to measure how well states are faring relative to each other, we calculate the difference between projected CFR’s for each state and the average CFR for the US (Fig 6). Positive (negative) values indicate that the CFR is worse (better). It clearly shows that there is a wide disparity between states’ case fatality rates. Furthermore, the difference in CFR closely corresponds with projected cumulative deaths. This shows even more dramatically how much greater New York’s and Connecticut’s outbreaks are compared to other states.

**Fig 6.**
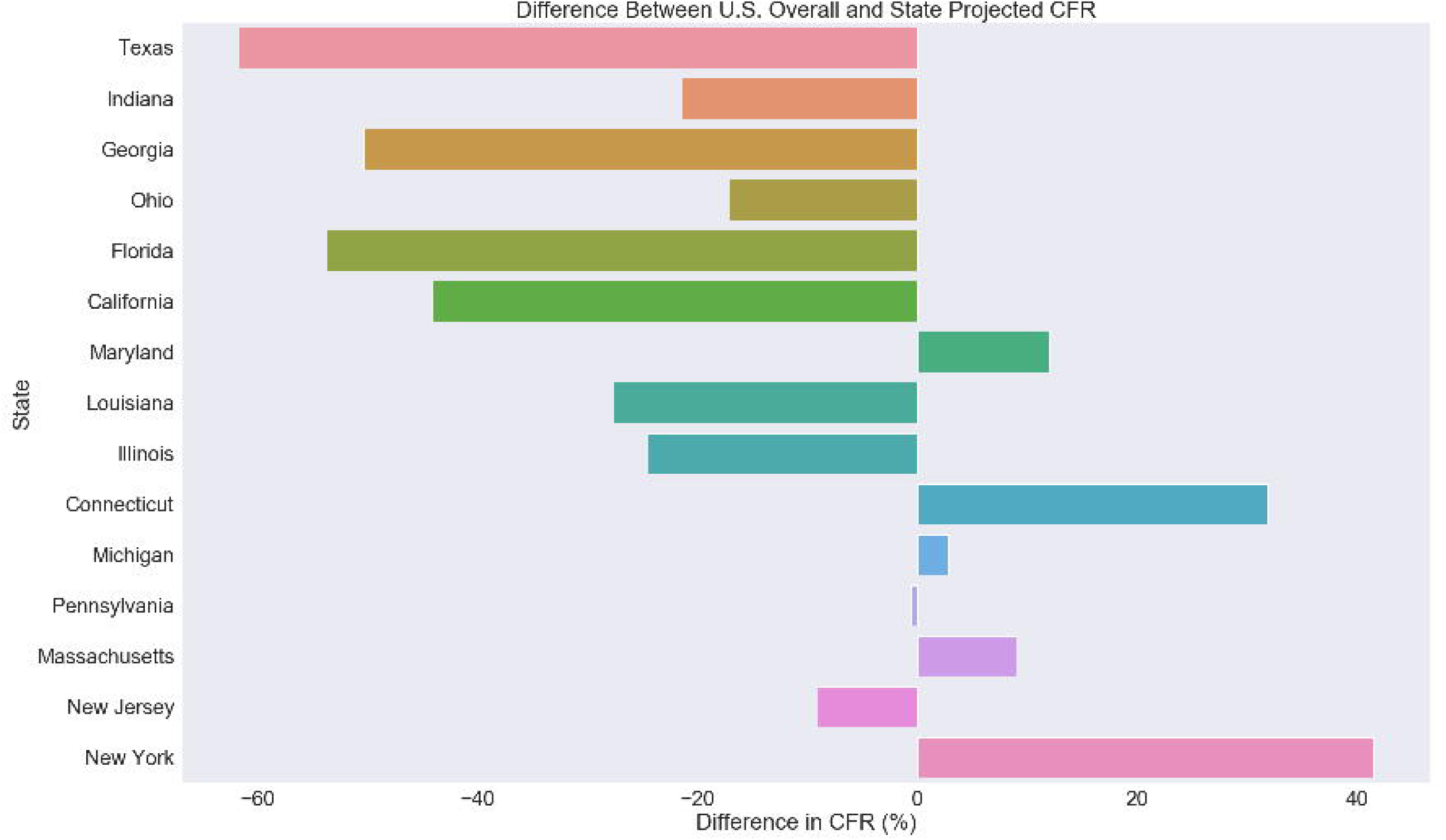
Difference in forecasted case fatality rates between states. Difference in forecasted case fatality rates between the top states that contributed 90% of all the deaths. This was calculated by subtracting the difference between projected CFR for individual states and the U.S average CFR.

### Predicting deaths

Considering that CFR and case incidence are the factors of death, it is fitting now to discuss the projected incidence of deaths (Fig 7). We calculated the number of deaths for each of the top 15 states that contribute 90% of the deaths. This reveals the tremendous disparity between the size of outbreaks in different states (see also S4 Table). The majority of states will experience less than 10,000 deaths. New York is once again a significant outlier; the model returns a minimum of 40,000 deaths and a mode of 65,000 deaths, 45% of the U.S. total and more than the next five jurisdictions combined. This can be traced back to the state’s relatively high projected case fatality rate and case incidence. New Jersey, and Massachusetts are expected to follow New York, with 18,000, and 10,000 respective projected deaths. Many states are disproportionately represented; some are over-indexed (over-represented) in the national death toll while others are under-indexed. Differences in deaths among jurisdictions are ultimately indicative of differences in CFR and case incidence. Sources of variation include the chronology and success of mitigation efforts, the prevalence of testing, and the distribution of age [19] and comorbidities [20] within populations at risk of infection.

**Fig 7.**
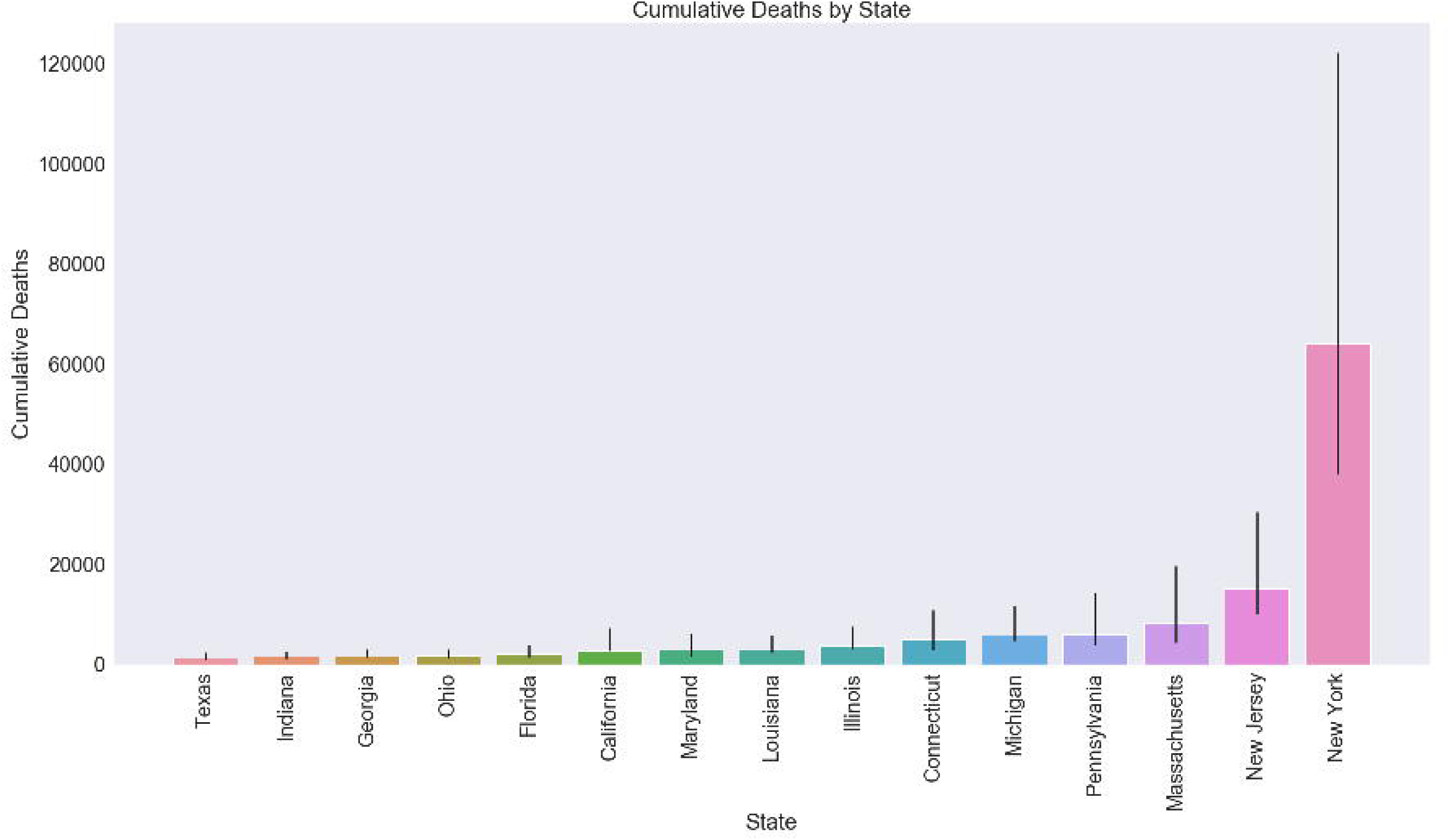
Forecasted deaths for the top US states. The range of forecasted deaths for the top states that contribute 90% of all the deaths. The low, mode, and high cases are displayed. The low and high cases are determined as the low and high end of the 95% confidence interval.

Table 1 summarizes the various possible death tolls under each of the 9 conditions. There is a 95% likelihood that any of these results are possible. The lowest cumulative deaths the U.S. could experience is 78,000, considerably higher than both Fauci’s prediction and the University of Washington model. The highest mortality is a sobering figure: 245000.

**Table 1.** Predicted death tolls for the US

|  | Low CFR | Mode CFR | High CFR |
| --- | --- | --- | --- |
| Low Case Incidence | 78833 | 97875 | 145282 |
| Mode Case Incidence | 100940 | 125484 | 183998 |
| High Case Incidence | 136079 | 168356 | 245904 |

Predicted deaths tolls for the nine different scenarios. These scenarios are constructed using the low, mode, and high cases for CFR and cases. The low and high cases are determined as the low and high end of the 95% confidence interval.

For the best-fit case in all the states, we calculated the number of deaths per day and the cumulative number of deaths (Fig 8). This shows that the number of deaths will reach an asymptote in the end of June and the number of daily deaths will peak in early May.

**Fig 8.**
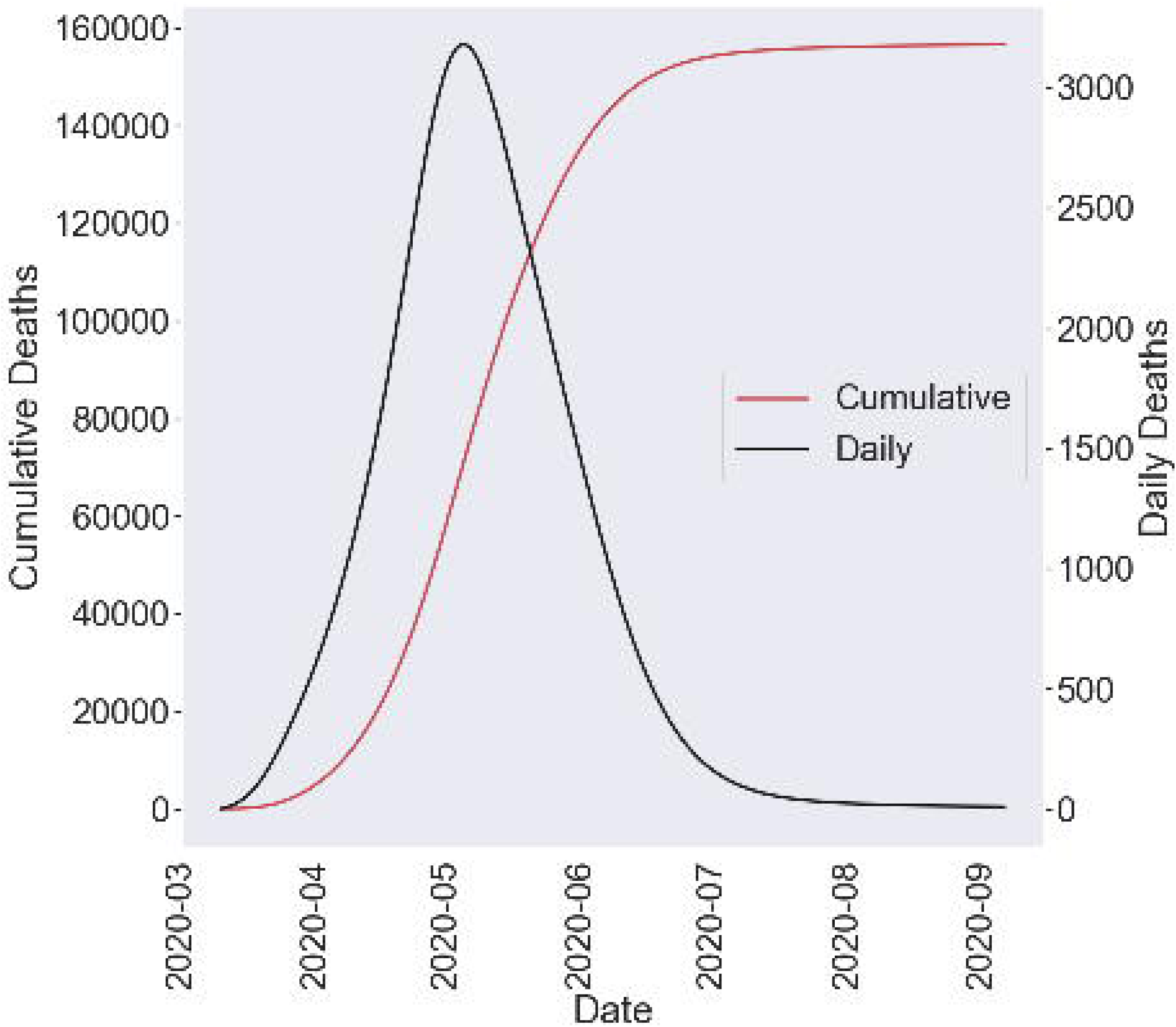
Forecasted deaths for US over time. The forecasted deaths of the United States over time. This shows both the new and cumulative number of cases.

## Discussion

We used a cohort analysis approach to estimate CFR and a modified logistic model (that explicitly accounts for the impact of mitigation efforts) to forecast case incidence on the state level, and afterwards calculated mortality on the state and national levels. Our model showed a wide range of mortality, with 79,000 deaths on the low end and a maximum of 245,000 deaths. Every possibility predicted by the model exceeds the prognostications produced by both the White House and the University of Washington model. Our model also revealed the deep disparity in deaths among different states, which is attributable to differences in case fatality rate and case incidence. We postulate reasons for these variations.

Many states in the US Northeast, including New York, New Jersey, Massachusetts, and Connecticut are disproportionately represented in the cumulative death toll. This disparity is primarily because these states have much worse case incidence and case fatality rates. New York is forecasted to experience the largest outbreak, the greatest CFR, and the highest mortality of any state by far. One explanation for its high case fatality rate is the strain the epidemic has placed on its healthcare system. As a result of its high case incidence, more hospitalizations will be required, overwhelming the medical care system. This could result in diminished quality of medical care, resulting in a high case fatality rate. Additionally, the disease has disproportionately impacted low-income, more vulnerable areas. [21]

The reason for the high case incidence itself is more perplexing. Numerous factors are likely at play, such as the popularity of public transportation [22] and the high population density of the New York City metropolitan area [23]] (where the vast majority of cases have been reported [24]). However, it is difficult to find conclusive evidence that any of these factors are directly accountable for the outbreak. It is very likely that luck played a large role in determining where clusters appeared. There is ample evidence that super-spreading events, or SSE’s, can cause sizable outbreaks [25]. For instance, officials in New York stated that as many as fifty infections could be traced back to a single man in Westchester County [26].

We propose two principal explanations for the discrepancy between the death tolls forecasted by the University of Washington model and that of our model: the differences in both the procedure for calculating CFR and the procedure for calculating mortality. Our cohort-based method to determining CFR’s predicts case fatality rates more accurately at every stage of the outbreak than other models because it explicitly accounts for the onset-to-death interval. Further, we forecast cumulative mortality by independently evaluating CFR and case incidence. In contrast, the University of Washington model directly predicts deaths; this method is prone to greater errors.

While both the CFR and case incidence models fit the data extremely well, there are several challenges with estimating the number of deaths accurately. Our model assumes the scale and methods of surveillance do not significantly change between today and the future. Any changes to the testing process will affect the number of confirmed cases. Breakthroughs in leveraging telemedicine, for instance, would result in increased detection of infected individuals. In this case, the model’s current forecasts for case incidence would be underestimates [27]. Additionally, if the shelter-in-place order is withdrawn from states too early, there will likely be an increase in both the case incidence and the mortality. The model itself has limitations; if our assumptions do not hold true, then our analysis will not hold true either.

## Supporting information

**S1 Table Goodness of fit** Goodness of fit for both the CFR and case incidence model. This shows excellent fit between the model and data.

**S2 Table Final cumulative case incidence for all states**. The predicted cumulative case incidence for all states shows significant variability across states.

**S3 Table CFR for all states**. The predicted CFR for all states shows significant variability across states.

**S4 Table Final number of deaths for all states**. The predicted deaths for all states shows significant variability across states.

## Data Availability

All data on COVID-19 are available from: https://github.com/CSSEGISandData/COVID-19
All data on United States state populations are available from: https://www.census.gov/data/datasets/time-series/demo/popest/2010s-state-

## Acknowledgments

I would like to thank Chandra Narayanan for all his guidance on building the best case incidence model.

---

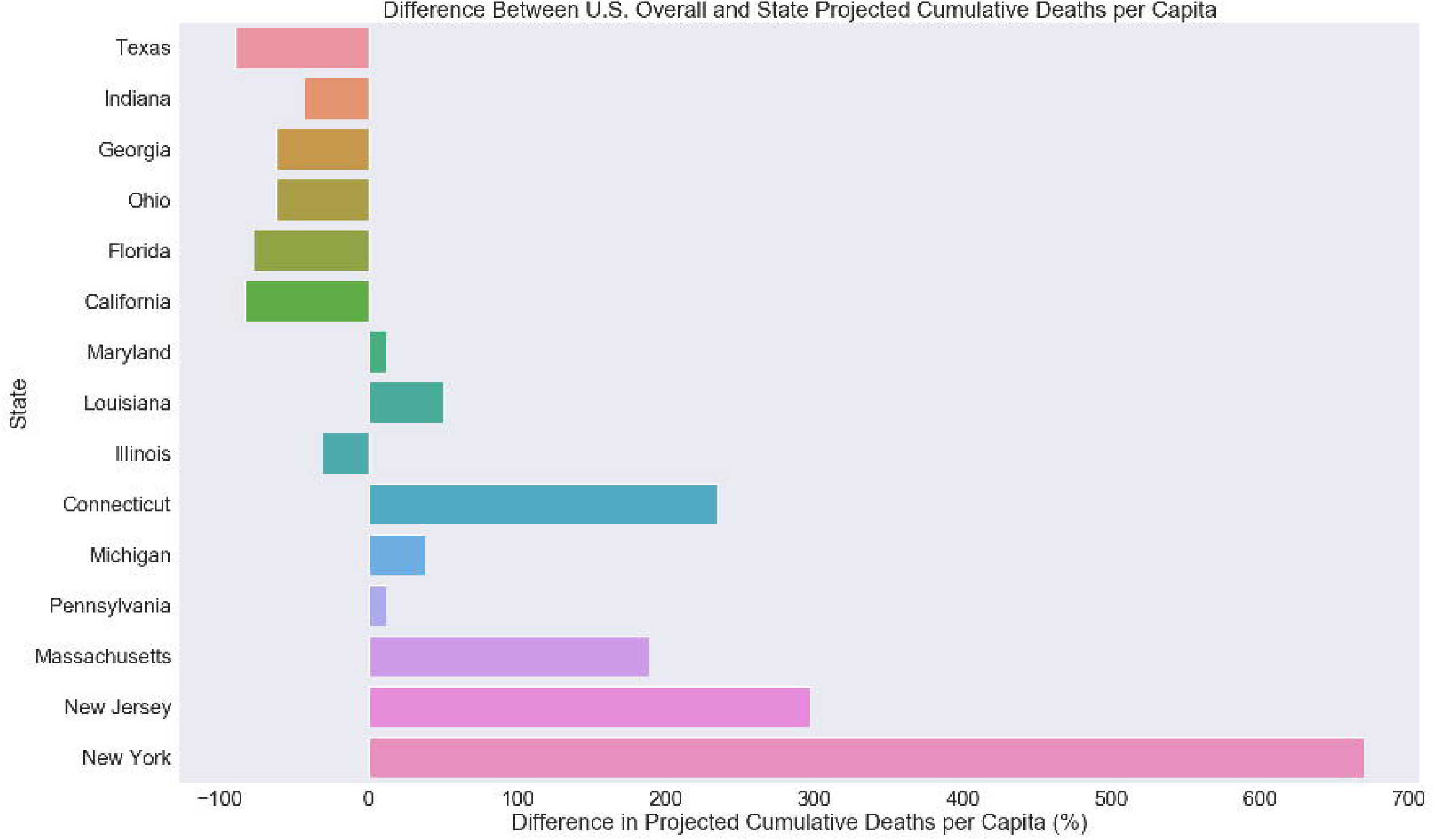

